# Hosting Displaced Medical Students in Times of Crisis: A Multi-National Qualitative Study Advancing the Consolidated Framework for Implementation Research (CFIR)

**DOI:** 10.64898/2026.07.09.26357620

**Authors:** Morteza Rezaei-Zadeh, Yaser Hamam, Shameq Sayeed, Simon Gay, Maha AbuZarifa, Khaled Zaqout, Ola AbuOlwan, Lian Massri, Lana Alhennawi, Farah Miqdad, Mohamed Zughbur

## Abstract

Catastrophic geopolitical conflicts increasingly disrupt the continuity of global medical education, placing immense pressure on clinical training pipelines and forced-migration student groups. While short-term, reactive, remote learning models exist, there are a profound lack of evidence-based implementation templates for medical schools within stable host nations to systematically host and integrate displaced clinical student cohorts mid-stream. This study explores the multi-level barriers and facilitators to hosting displaced medical students across diverse international environments, seeking to establish a rigorous, scalable model of educational sanctuary while advancing implementation science theory in crisis contexts. Employing a qualitative multi-site case study design guided by a critical realist ontology, this study analysed 66 semi-structured interviews with displaced Gazan medical students, hosting lecturers, clinical coordinators, and support staff across the United Kingdom, Malaysia, Pakistan, Turkey, and South Africa, mapping reflexive thematic analysis findings onto the Consolidated Framework for Implementation Research (CFIR).

The analysis revealed that while rigid immigration policies, clinical placement caps, and severe cultural distance represent substantial barriers, key facilitators include assessment considerations, flexible placement models, sanctuary institutional cultures, peer networks, and decentralised administrative trust. Strategic administrative approaches, such as classifying displaced students as extended clinical elective visitors rather than full-time matriculants, enabled institutions to accommodate them within existing frameworks.

This study demonstrates that public sector higher education institutions can act as vital global sanctuary networks to preserve clinical training pipelines. Crucially, the findings advance implementation science by proposing three novel constructs for the updated CFIR in crisis environments: Agile Implementation Over Perfection within the Implementation Process domain, Protective Leadership Shielding within the Inner Setting domain, and Bidirectional Boundary Subversion at the Inner/Outer Setting interface. This theoretical refinement transforms CFIR from a determinant model for stable, clinical interventions into an active, equity-driven framework for rapid humanitarian response in politically contested environments.

## Introduction

The continuity of global healthcare training is increasingly threatened by high-intensity geopolitical conflicts, natural disasters, and systemic persecution that trigger unprecedented levels of forced migration and institutional collapse [1]. Forcible displacement has reached historic proportions, with current global estimates indicating that over 122.6 million individuals are actively fleeing persecution, war, and structural violence [2]. Higher education has historically remained a highly restricted privilege for displaced populations; despite education being a fundamental human right, only approximately one per cent of refugee and displaced youth globally manage to access or continue higher education, with the vast majority forced to abandon their academic endeavours [4]. In the field of medical training, this educational barrier is exacerbated by highly complex professional regulations, national clinical placement caps, and stringent licensing standards [3].

Geopolitical conflicts in regions such as Afghanistan, Gaza, Sudan, Syria, Ukraine, and Yemen have not only displaced thousands of aspiring healthcare professionals but have also physically devastated local training environments, clinical simulation laboratories, and teaching hospitals [1]. For instance, in Sudan, the ongoing armed conflict has led to the displacement of over 10 million people and the operational collapse of more than two-thirds of the country’s healthcare and educational facilities, with approximately 79.3% of dental and medical schools experiencing direct military assaults and looting [5]. In Ukraine, the recent conflict has severely disrupted medical education standards, destroying crucial clinical infrastructure and forcing both domestic and international medical students to seek education across Europe [6]. Similarly, in Syria, half of the 31,000 reported physicians were displaced, and by 2013, 70% of the health workforce had fled the country, severely compromising post-conflict healthcare rebuilding efforts [2]. In Gaza, high-intensity military actions have resulted in the widespread destruction of academic infrastructure, creating an environment of ‘scholasticide’ where the entire physical and administrative apparatus of medical training is systematically dismantled [23].

Despite these compounding crises, the literature surrounding displaced students in higher education has historically been limited and fragmented. The primary focus of existing research has been confined to compulsory primary and secondary education [7], general international student acculturation [9], or individual-level psychological trauma and acculturative stress [9]. Within the clinical training domain, studies often examine either the short-term, reactive provision of online distance learning [1] or the post-hoc labour market integration of fully qualified, foreign-trained immigrant health professionals [10]. A profound empirical gap exists regarding how medical schools within stable host nations can systematically organise, design, and implement formal hosting programmes that temporarily or permanently integrate displaced medical students into mid-stream clinical cohorts [2]. Medical education operates under exceptionally rigid structural constraints; unlike general humanities or social science disciplines where curriculum mapping is flexible and regulatory oversight is minimal, clinical training requires strict alignment with national medical councils, patient safety protocols, and hospital capacity limits. Furthermore, although implementation science has produced robust, multi-level determinant frameworks - most notably the Consolidated Framework for Implementation Research (CFIR) - their application has historically been limited to stable clinical environments or the introduction of evidence-based health-service innovations [12]. There is a complete lack of research utilising and adapting such frameworks to evaluate the implementation of rapid, crisis-driven humanitarian interventions within highly regulated public sector educational institutions [15].

This study directly addresses these critical gaps by conducting an exhaustive, multi-national qualitative analysis of the barriers and facilitators to hosting displaced medical students. Based on a robust dataset representing 66 in-depth interviews with displaced students, hosting academic lecturers, clinical coordinators, and university support staff operating across diverse international host contexts, including the Malaysia, Pakistan, South Africa, Turkey, and United Kingdom, this paper systematically evaluates the structural, cultural, and political dynamics of institutional integration. By framing this analysis through the lens of the Consolidated Framework for Implementation Research (CFIR) [25], the study identifies key operational determinants and, crucially, advances the theoretical boundaries of CFIR to accommodate highly dynamic, politically charged, and crisis-driven public sector environments.

To provide a structured and comprehensive analysis, this research inquiry is organised around five central research questions:

1. What features of the hosting programme, such as cost and academic fit, affect the school’s decision to participate?
2. What external laws, policies, and social pressures influence the school’s willingness to host?
3. What internal resources and aspects of school culture act as barriers or facilitators?
4. What attitudes and beliefs of staff and faculty impact the acceptance of the programme?
5. What planning and leadership factors help or hinder the successful start of the programme?

### Theoretical Framework

To systematically evaluate the multi-level contextual determinants and individual agency within this public sector intervention, this study utilises the updated (2022) Consolidated Framework for Implementation Research (CFIR) [13]. CFIR operates as a meta-theoretical determinant taxonomy that identifies the real-world barriers and facilitators influencing programme effectiveness [12, 13]. The framework evaluates implementation across five interactive domains, operationalised here as:

- **The Innovation:** The intrinsic characteristics of the educational hosting programme, including its complexity, cost, adaptability, and relative advantage over alternative pathways [11, 16].
- **The Outer Setting:** The external sociopolitical, legal, and regulatory context, encompassing national immigration laws and educational regulatory policies [13].
- **The Inner Setting:** The internal structural and cultural characteristics of the hosting medical school, evaluating its pre-existing ethos, infrastructure, and leadership readiness [13].
- **The Individuals:** The roles, beliefs, and attributes of the actors involved, including institutional deliverers (faculty and coordinators) and recipients (domestic and displaced students) [12].
- **The Implementation Process:** The strategic phases, adaptive manoeuvres, and leadership actions used to plan, execute, and evaluate the initiative [14, 16].

CFIR is uniquely suited to this study for three reasons. First, hosting displaced students is a multi-systemic intervention; CFIR allows researchers to systematically map intersecting levels - from national visas to local university execution - without overlooking their causal links [13]. Second, the 2022 CFIR update centres innovation recipients and equity [14], ensuring the unique vulnerabilities and structural barriers faced by displaced students remain central to the analysis [8]. Finally, CFIR provides operationalised qualitative tools [13] that ensure systematic comparability and methodological rigor across highly diverse host nations like the UK, Pakistan, and South Africa [15].

### Methodology

To systematically evaluate the complex, multi-level factors that influence the implementation of hosting programmes for displaced medical students, this study utilises a rigorous, contextually sensitive qualitative research design. The methodology is structured across six key dimensions, explaining not only the execution of each phase but also the theoretical and practical justifications for each chosen strategy.

### Philosophical Underpinning

This study is rooted in the philosophical paradigm of Critical Realism (CR). Critical realism provides a highly sophisticated ontology that is suited for health services, policy, and educational implementation research [18]. It posits that reality is stratified into three distinct, non-congruent domains:

1. **The Empirical**: The surface-level events, observations, and direct experiences that actors perceive.
2. **The Actual**: Events and behaviours that occur in the world, regardless of whether they are directly observed or recorded.
3. **The Real**: The deep, underlying structural mechanisms, causal powers, and generative structures that possess the capacity to produce actual events [18].

Within a critical realist framework, causal mechanisms are understood not as rigid, linear laws, but as *tendencies* that interact dynamically with human agency and local contexts to produce complex, sometimes unpredictable outcomes [18]. This philosophy is exceptionally appropriate for this study because the integration of displaced medical students is a highly contested social and institutional process. A naive positivist approach would fail by attempting to reduce implementation outcomes to simplistic, linear variables (such as funding or test scores) [19], while an extreme relativist or social constructivist approach would fail by neglecting the very real, hard structural constraints of national borders, immigration laws, and clinical capacity caps. Critical realism allows this study to recognise the objective reality of these external, rigid structures (the Real) while simultaneously honouring the subjective agency and moral strivings of institutional actors and students who work to subvert or adapt within those structures to facilitate educational continuity [18].

### Study Design

The study employs a Qualitative Multi-Site Case Study Design. A qualitative approach is most appropriate when the primary goal is to understand complex, contemporary social phenomena within their real-world contexts, particularly where the boundaries between the phenomenon and the context are highly blurred [12]. Given that the hosting initiative was implemented across vastly different geopolitical, legal, and cultural environments (ranging from high-resource, de-centralised higher education systems in the UK to highly structured clinical hierarchies in Malaysia and Pakistan), a multi-site case study design enables the research team to explore localised demi-regularities - patterns of barriers and facilitators that occur under specific contextual conditions [19]. This design ensures that the unique socio-ecological conditions of each host site are fully preserved and analysed, rather than being averaged out into descriptive statistics.

## Method

The primary data collection method consisted of Semi-Structured Qualitative Interviews. Semi-structured interviews are appropriate because they employ a standardised interview guide mapped directly onto the core domains of the theoretical framework (CFIR), ensuring high systematic comparability across the diverse multi-national dataset [12]. Concurrently, they offer the flexibility required to probe deeply into participants’ subjective experiences, emotional negotiations, and localised structural navigations [20]. This approach allowed participants, particularly vulnerable displaced students, to articulate their realities in their own words, revealing unanticipated barriers and organic coping strategies that would have been completely obscured by rigid, structured surveys [17].

### Population and Sampling

This study targeted a highly diverse, multi-stakeholder population involved in the design, administration, delivery, and lived experience of the hosting initiative. The target population comprised:

- Displaced medical Gazan students who were hosted at various international sites (UK: n=2, Egypt: n=16, Malaysia: n=8, Pakistan: n=9, South Africa: n=9, Turkey: n=8).
- Academic lecturers and clinical tutors who delivered pedagogical content (n=6).
- Clinical placement coordinators and directors who managed hospital rotations (n=4).
- University administrative and support staff (including IT, accommodation, and sanctuary unit coordinators, n=4).

Participants were selected using purposive sampling based on their direct experience with the phenomenon under study. Snowball sampling was utilised as a secondary strategy to recruit displaced medical students; leveraging trusted peer networks was essential to navigate institutional reticence among this legally precarious population [2] and ensure equitable sample representation [17]. The final sample size was 66 in-depth interviews distributed across five international contexts: the UK, Malaysia, Pakistan, Turkey, and South Africa. The transcribed interviews produced a total of 451 pages of interview transcripts, comprising 115,154 words.

### Data Gathering and Analysis

Interviews were conducted either in person or via secure, encrypted video-conferencing platforms (due to geographical dispersion and safety considerations), and were fully transcribed verbatim. The dataset was analysed using Reflexive Thematic Analysis (RTA), following the rigorous six-phase recursive framework developed by Braun and Clarke [20]:

1. *Familiarisation*: Repeated reading of transcripts and generating initial visual mind-maps.
2. *Generating Codes*: Systematic, line-by-line coding of the entire dataset, employing both an inductive approach (to capture emergent personal experiences) and a deductive approach (mapping codes to CFIR constructs).
3. *Constructing Themes*: Collapsing codes into broader, multi-level thematic patterns.
4. *Revising Themes*: Reviewing candidate themes against the coded extracts and the entire dataset to ensure coherence.
5. *Defining Themes*: Refining the specific conceptual boundaries of each theme and sub-theme.
6. *Producing the Report*: Writing the analytical narrative, weaving in dense theoretical interpretations and direct participant quotes [20].

Reflexive thematic analysis is uniquely appropriate for this study because it explicitly rejects the notion of the researcher as a passive, value-free instrument, instead positioning the researcher’s critical subjectivity as a vital tool for deep interpretive analysis [18]. This aligns perfectly with a critical realist philosophy, which acknowledges that while structural mechanisms exist objectively, our knowledge of them is always socially produced and interpretively mediated [18].

### Trustworthiness

To ensure the highest academic standards of qualitative rigor, this study adhered strictly to Lincoln and Guba’s established criteria for trustworthiness [24]:

- **Credibility**: Established through researcher and data source triangulation. Multiple members of the research team independently coded a subset of transcripts, and coding discrepancies were resolved through reflexive team meetings to ensure conceptual alignment [20]. Furthermore, findings were triangulated across different participant groups (students vs. deans) to ensure a balanced multi-perspective account [12].
- **Transferability**: Achieved by providing thick, detailed contextual descriptions of the host countries’ healthcare, legal, and academic structures, enabling future researchers to assess the applicability of these findings to other institutional settings [14].
- **Dependability**: Ensured by maintaining a meticulous, transparent audit trail documenting all analytical decisions, raw coding frames, and theme development iterations [20].
- **Confirmability**: Maintained through continuous reflexive journaling by the researchers to identify and mitigate personal preconceptions, and by ensuring that every qualitative claim is directly supported by extensive verbatim participant quotes, rendering the analysis highly transparent and grounded in the raw data [21].

The study received ethical approval and the written consent was received prior to participants’ participation in the study. The start date of the recruitment was 22 June 2025 and the end date was 12 Dec 2025.

## Findings

The structural and qualitative findings of this multi-national study are organised and presented sequentially under the five core research questions. For each research question, a comprehensive Markdown table summarises the themes, conceptual definitions, corresponding empirical codes, and source interviews, followed by an in-depth, continuous qualitative interpretation that integrates direct participant quotes.

### Research Question 1: Features of the Hosting Programme Affecting Participation

This research question examines what specific features of the hosting programme - encompassing structural, academic, financial, and technological elements - influence a medical school’s decision to participate in hosting displaced medical students.

Each theme generated from the analysis and presented in Table 1 is further interpreted below.

**Table 1.** Features of the Hosting Programme Affecting Participation in Hosting Displaced Students.

| Theme | Theme Definition | Corresponding Concepts | Interview No. |
| --- | --- | --- | --- |
| **Formative Assessment Adaptation** | Utilising formative rather than summative assessments to address local degree-awarding risks while providing academic evidence. | Formative OSCEs, Non-summative risk mitigation, Progression decision decoupling | Int 2, Int 8, Int 10, Int 14 |
| **Dual-Curriculum Navigation** | The programmatic allowance for students to simultaneously satisfy host requirements and original home school mandates. | Dual-curriculum balancing, Flexible attendance tracking, Home school online exams | Int 2, Int 3, Int 6, Int 13 |
| **Clinical Competence Equivalency** | The degree to which prior medical training translates into the host country's clinical expectations. | Baseline competence assumptions, Clinical rotation integration, OSCE unfamiliarity | Int 8, Int 1-4, Int 2-4, Int 4-8 |
| **Bespoke Technological Provision** | Providing specific technological tools and digital orientations to ensure academic parity with local cohorts. | Blackboard VLE access, iPad distribution, Learning technologist support | Int 2, Int 13, Int 15 |
| **Asymmetric Academic Mapping** | The challenge of mapping disparate curriculum structures, particularly regarding early clinical | Curriculum misalignment, Grade translation discrepancies, Pre-clinical vs | Int 2, Int 7, Int 13 |
|  | exposure. | clinical mismatch |  |
| **Pedagogical**<br>**Shift in**<br>**Content**<br>**Delivery** | The structural adjustments in how lectures and clinical skills are taught to a displaced cohort. | Online distance learning challenges, Reliance on AI tools, Absence of physical labs, Student-led presentation models, Loss of clinical exposure | Int 6-1,<br>Int 6-4,<br>Int 6-8,<br>Int 2-4 |

#### 1. Formative Assessment Adaptation

A major programmatic barrier identified was the risk associated with summative assessments; faculty feared that assessing displaced students formally could result in academic failure or regulatory complications. To facilitate participation, the programme adapted its assessment strategy. As one assessment lead recalled regarding past failures with transferred international students, “*I felt quite strongly that they shouldn’t be taking our summative examinations and that we shouldn’t be determining their progression”*. The facilitator was the shift to formative assessment. Students took exams *“formatively, so that means just to give them feedback… that could then be used by their own universities to inform progression decisions”*. This feature decoupled the host institution from the legal and academic risk of awarding a degree, thereby drastically increasing the school’s willingness to host.

#### 2. Dual-Curriculum Navigation

The hosting programme’s structure necessitated a feature that allowed students to navigate two distinct educational realities simultaneously, which acted as both a barrier and a facilitator. Students reported intense pressure attempting to balance the host’s clinical placements with their home institution’s online requirements. One tutor observed the students “*having to trade two curriculum at the same time,”* managing *“monthly modules and passing those modules at the home school”* while attending GP placements in the UK. One displaced student admitted, “*I couldn’t balance Leicester’s studies alongside with my university studies, so I had eventually to take my own decision about not doing Leicester’s exams”.* The programme’s flexibility, allowing students to take time off host placements to revise for home exams, was a vital facilitator. Without this programmatic flexibility, the initiative would have forced students to abandon their original degrees, ultimately undermining the goal of institutional continuity.

#### 3. Clinical Competence Equivalency

The perceived clinical readiness of displaced students significantly influenced the school’s decision to integrate them into patient-facing environments. The assumption of foundational competence was required to allow students into wards. A senior faculty member noted, *“we had to assume that their intelligence and attitude was equivalent to medical students in this country”*. However, the reality of clinical equivalence varied. In some host contexts, students noted that while their extensive theoretical knowledge was praised, they lacked the specific practical and communication skills required by localised assessment formats like OSCEs. A student reflected, *“when I came to the OSCE for the first time, that was a shock for me that I’ve never done those before”.* In South Africa, a student noted, “*my instincts were for someone else to do the hands on… In South Africa, being hands on was very demanded and I had to learn how to be more active”.* A programmatic feature that facilitated integration was allowing students to observe and gradually participate without the immediate burden of clinical autonomy.

#### 4. Bespoke Technological Provision

To ensure the displaced students could effectively engage with the host curriculum, the programme featured bespoke technological provisioning. Digital inequity was recognised as an immediate barrier. The programme facilitated inclusion by mirroring the technological resources given to first-year students. An IT coordinator noted the necessity of ensuring *“the technology that we get works on the iPad or on some other device… and also things like logins, accounts”.* Students were *“issued with an iPad”* and *“put… in touch with our learning technologist who could teach them how best to use that”.* This feature not only removed logistical barriers to the curriculum but actively facilitated the students’ psychological integration, signalling that they were valued and equipped equivalently to their local peers.

#### 5. Asymmetric Academic Mapping

The inherent asymmetry between the curriculum of the host nation and the conflict-zone universities presented a complex programmatic barrier. Grade translation and course equivalency required intense negotiation. A coordinator from Gaza highlighted that *“a 70 in the University of Leicester maybe means 90 in the Islamic University… the marking criteria or the numbers doesn’t reflect what is marked exactly in the original university”*. The host programme facilitated solutions by providing raw data and formative feedback directly to the home deans, allowing the home institutions to “*accredit that learning to be equivalent to what their requirements would be”.* This feature respected the autonomy of the original institutions while allowing the host school to avoid the bureaucratic challenge of formal credit transfer.

#### 6. Pedagogical Shift in Content Delivery

A critical programmatic feature that acted as a barrier for many students, particularly those shifted to fully remote models in Egypt, was the abrupt change in how medical content was delivered. Students highlighted the devastating loss of physical anatomy labs and direct faculty interaction. One student lamented, *“our education became based on theory only. We do not have any labs that can apply the information… when I studied it, I struggled a lot to understand the body and anatomy properly”.* To cope with this barrier, students engaged in sophisticated self-directed learning, relying heavily on third-party tools. As one student noted, *“artificial intelligence (AI) had a very big importance in this matter… Google or the internet in general feels like the first helper for students at this time”.* The programme’s reliance on self-directed online modules forced a severe pedagogical shift, challenging students’ confidence but ultimately building extreme academic resilience.

### Research Question 2: External Laws, Policies, and Social Pressures

This research question explores the macro-level external forces - including national immigration frameworks, professional medical councils, governmental stances, and societal solidarity movements - that influence a medical school’s willingness and capacity to host displaced medical students.

Each theme generated from the analysis and presented in Table 2 is further interpreted below.

**Table 2.**
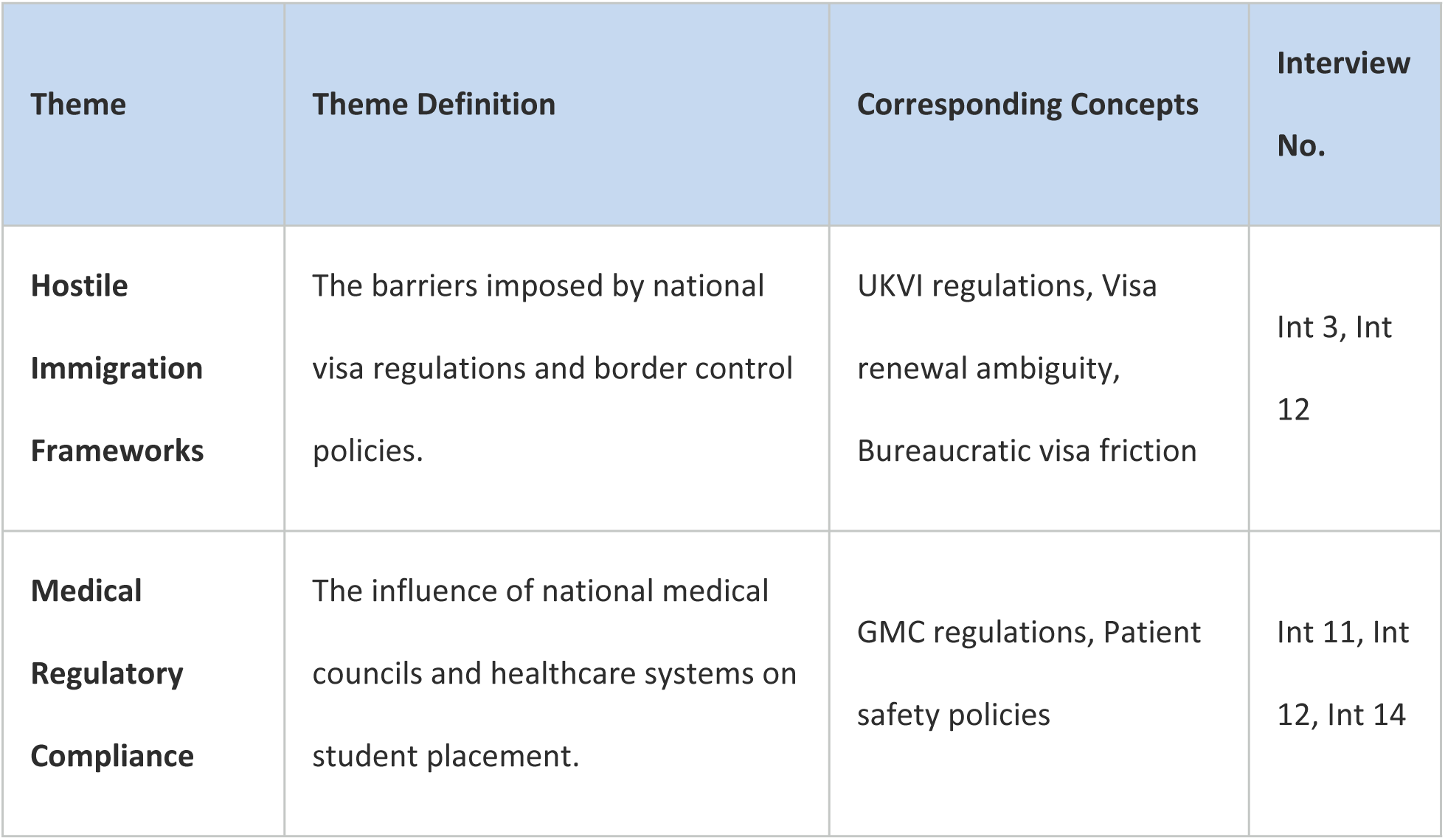
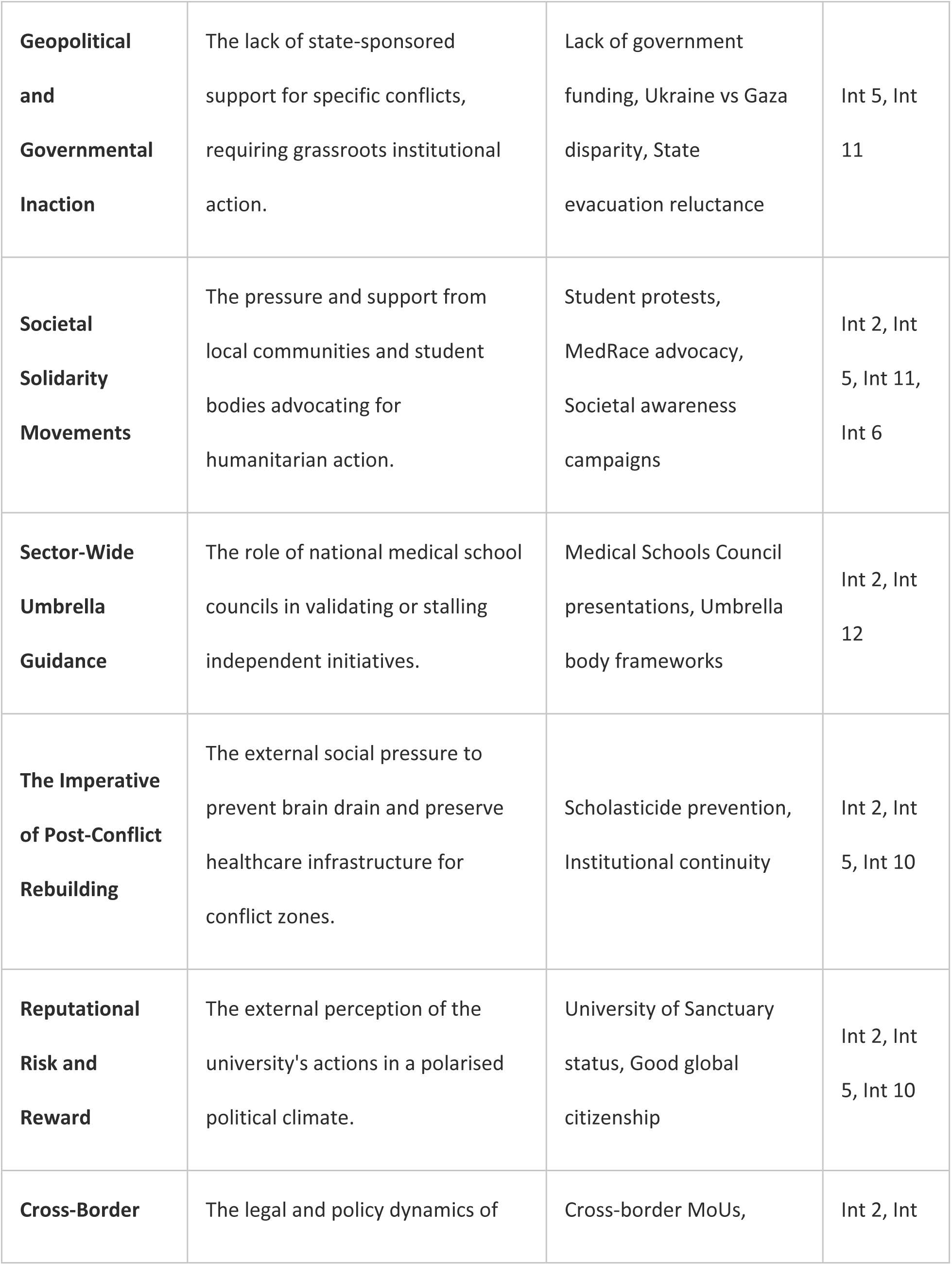
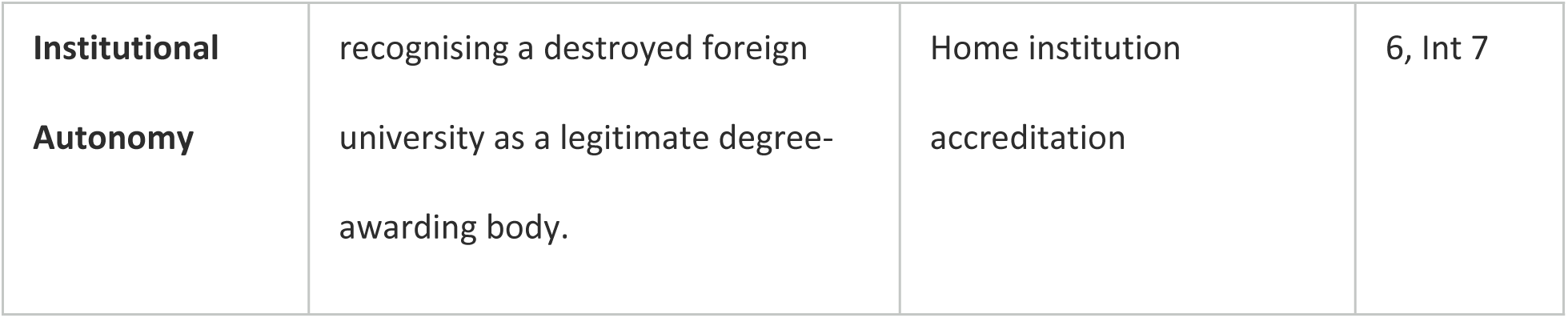
Legal, Policy, and Social Factors Affecting Participation in Hosting Displaced Medical Students.

#### 1. Hostile Immigration Frameworks

National immigration policies operate as the most severe external barrier to hosting displaced students. The data reveals intense friction between the humanitarian goals of the medical school and the rigidity of border control agencies. One senior academic bluntly characterised the national immigration stance as a hostile environment, noting that the “*UKVI… assumption is the student is bad and we won’t give a visa rather than the other way”*. This systemic resistance manifested directly in the students’ lives, with personal tutors reporting students facing “*conflicting advice from different sections in terms of what do they put in their visa application for renewal*”. The ambiguity between holding a visitor visa versus a student visa while conducting extended electives forced the institution to navigate perilous legal grey areas, heavily influencing the scale and speed at which they could confidently accept students.

#### 2. Medical Regulatory Compliance

In medical education, external healthcare providers and regulatory bodies hold immense sway over student placements. The willingness of a university to host can be contingent upon the cooperation of organisations like the NHS and the General Medical Council (GMC). A facilitator in hosting displaced students was the willingness of local NHS trusts to accept the students using already established elective processes without making additional demands from this already disadvantaged group. A university leader confirmed making *“partner NHS trusts aware of this initiative… and I’ve had universal positivity about it*”. However, the shadow of regulatory compliance remained a barrier; as one dean noted, navigating “*NHS regulations that seek to satisfy patient safety*” alongside GMC regulation created a web of stakeholders whose rules could easily obstruct the initiative if misaligned. The programme survived by integrating students under the existing liability and educational cover of local cohorts.

#### 3. Geopolitical and Governmental Inaction

The medical school’s willingness to host was paradoxically catalysed by a profound lack of external governmental support. Interviewees repeatedly contrasted the robust state-sponsored mechanisms created for Ukrainian refugees with the glaring silence regarding Gaza. One UK colleague pointed out that “*in the context of the Ukraine conflict, the government pulled out all the stops… In the case of Gaza, the government has done very, very little*”. This external inaction acted as a powerful social pressure on the university to step into the void. Faculty viewed the initiative as a *“powerful way that universities can demonstrate to the government… that we can find ways ourselves to do something… because it’s the right thing to do”*. Thus, state-level apathy served as a moral facilitator, pushing the institution to construct independent pathways.

#### 4. Societal Solidarity Movements

Internal and external societal pressures significantly facilitated the school’s commitment to the initiative. There was a palpable student voice pushing back against the genocide and demanding institutional accountability. Grassroots groups like MedRACE (https://le.ac.uk/cls/cls-equality/medrace) and the University of Leicester Palestine Society applied constructive pressure by organising bake sales, charity dinners, and exhibitions that raised substantial funds. This internal societal solidarity proved to leadership that the initiative had broad community backing. As one senior leader observed*, “society needs a good dose of that from time to time to remind itself of what’s important”*. The visible support from the student body and local community effectively neutralised concerns that the initiative might be divisive, validating the executive board’s decision to proceed.

#### 5. Sector-Wide Umbrella Guidance

External policy bodies, specifically the Medical Schools Council (MSC), played a dual role as both an initial barrier and an eventual facilitator for hosting the medical displaced students. Initially, the lack of unified national guidance meant universities operated in silos, making the legal and logistical hurdles seem insurmountable. However, once the pilot of hosting the displaced students proved successful, the MSC became a facilitator for scaling the programme. Presentations to the MSC allowed the host institution to *“present it to all medical schools across the country to say here’s the initiative”.* This sector-wide networking helped establish a “*global network who are supported with the initiative,”* easing the perceived risk for individual schools by embedding their actions within a recognised, multi-institutional framework.

#### 6. The Imperative of Post-Conflict Rebuilding

A profound external social pressure driving the initiative was the recognised necessity to combat ‘scholasticide’ and preserve the healthcare pipeline for the conflict zone. The analysis reveals a deep understanding among faculty that simply extracting students permanently would constitute a brain drain contributing to the *“death of those schools themselves”*. The programme was fundamentally shaped by the external policy goal of *“rebuilding the healthcare and healthcare education infrastructure in Gaza”.* This pressure facilitated a model where students remained tethered to their original universities. By framing the initiative as an act of institutional preservation as well as individual rescue, the medical school aligned itself with global humanitarian objectives, thereby increasing institutional willingness to invest resources.

#### 7. Reputational Risk and Reward

The polarised political climate surrounding geopolitical conflicts generates significant external pressure. The university had to navigate the reputational risks associated with perceived partisan action. However, the institution’s existing external branding as a ‘University of Sanctuary’ facilitated a protective policy umbrella. A staff member noted that while there might be people in society *“who are not happy that these sorts of schemes… are happening, this provided no reason for us not to do it”.* In fact, acting in accordance with their sanctuary status enhanced the university’s reputation for social responsibility. The external pressure to live up to their stated civic values ultimately outweighed the fear of political backlash, driving the programme forward.

#### 8. Cross-Border Institutional Autonomy

The legal logistics of interacting with foreign universities operating under siege conditions presented unique external policy barriers. Standard cross-border higher education agreements (MoUs) require extensive legal vetting, which was impossible given the destruction of the home institutions. A facilitator was the willingness of both host and home deans to operate on trust and informal coordination rather than rigid legal contracts. A coordinator from Gaza explicitly noted, “*I’d rather organise this without a formal MoU with the university, as we have a colleague there who we know well and can rely on to help make things happen.”.* This reliance on agile, relationship-based coordination alongside existing policy and governance frameworks was essential to launching the initiative swiftly.

### Research Question 3: Internal Resources and Aspects of School Culture

This research question investigates how internal setting dynamics - including clinical training cultures, dedicated staff roles, peer networks, communication pathways, and financial resource models - operate as barriers or facilitators to hosting initiatives.

Each theme generated from the analysis and presented in Table 3 is further interpreted below.

**Table 3.** Internal Resource and School Culture Factors Affecting Participation in Hosting Displaced Medical Students.

| Theme | Theme Definition | Corresponding Concepts | Interview No. |
| --- | --- | --- | --- |
| **Egalitarian Student Integration** | The cultural commitment to treating displaced students identically to home students to foster belonging. | Anonymous integration, Embedded cohorts | Int 2, Int 11, Int 13, Int 14 |
| **Differentiated Pastoral Infrastructure** | The recognition that standard support systems may be inadequate for severe trauma and cultural displacement. | Psychological mismatch, Personal tutor limitations | Int 2, Int 4, Int 13 |
| **Cultural Translators and Link Tutors** | The deployment of specific faculty to act as bridges between the host and home curriculums and cultures. | Link Tutor role, Cross-system navigation | Int 2, Int 13 |
| **Sanctuary Institutional Ethos** | A preexisting cultural framework within the university that normalises and encourages refugee support. | Civic duty ethos, Inclusive learning environment | Int 2, Int 5, Int 10 |
| **Peer-to-Peer Support Systems** | Leveraging the student body to provide informal, social, and academic orientation. | Near-peer mentoring, Shared study spaces | Int 2, Int 12, Int 4-6 |
| **Ground-Level Communication Deficits** | The barrier created when high-level strategic plans fail to reach or be understood by frontline clinical staff. | Ignorant placement providers, Attendance misunderstandings | Int 3, Int 13 |
| **Flexible Placement Allocations** | The internal resource capacity to adapt clinical schedules to accommodate dual-curriculum demands. | Adapted placement models, Timetable modifications | Int 3, Int 11 |
| **Hierarchical Distance in Clinical Settings** | The cultural differences in student-doctor dynamics that create friction in host hospitals. | Student vs. doctor dynamics, Communication formality, Loss of university life, Extreme academic pressure | Int 2-6, Int 3-1, Int 3-2, Int 6-1 |
| **Financial Burden Mitigation** | The removal of direct educational and living costs for the student through institutional waivers. | Tuition fee waivers, Accommodation support, Cost-neutral enrolment | Int 1, Int 2, Int 4, Int 13 |
| **Philanthropic Capital Integration** | The reliance on external charitable organisations to sustain the programme without draining university budgets. | Living cost stipends, Third-party charity partnerships | Int 2, Int 4, Int 6, Int 14 |

#### 1. Egalitarian Student Integration

The prevailing culture of the medical school heavily favoured egalitarianism, integrating the displaced students anonymously into the standard cohorts. This cultural aspect acted as a major facilitator for the students’ sense of dignity. As one coordinator explained, the goal was to “*recreate that sense of identity and hopefully bring back some of that loss… being a medical student”.* Clinical placement leads corroborated this, stating, *“We treat them essentially as a Leicester student… we don’t know that they’re a displaced student… unless they volunteer that information”.* While this cultural blindness prevented stigmatisation and othering, it occasionally acted as a barrier when students silently struggled with unacknowledged knowledge gaps, demonstrating the dual-edged nature of egalitarian integration.

#### 2. Differentiated Pastoral Infrastructure

A significant barrier emerged regarding the internal resources dedicated to pastoral and academic support. While the university offered its standard counselling services, these were often culturally or practically mismatched to the students’ acute needs. Students reported feeling like a sample for the initiative rather than receiving targeted help, noting that personal tutors *“were not fully aware of my situation and therefore could not provide support as ideally they should”.* A student remarked that they had to explain myself every time because the standard personal tutoring infrastructure lacked the specialised context of dual-curriculum management and war trauma. This highlights that standard university pastoral resources are often inadequate for the complex psychological and academic realities of displaced scholars. Despite this, students were grateful to their personal tutors in supporting and signposting them, despite their differing needs to host School students.

#### 3. Cultural Translators and Link Tutors

To overcome the limitations of standard pastoral care, the introduction of a ‘Link Tutor’ served as a highly effective internal resource. Acknowledging that regular personal tutors only understood the UK system, the Link Tutor role was designed to understand *“both the home environment and the host environment well”*. This resource proved invaluable in mediating the friction between the two academic cultures. When a student felt overwhelmed by the expectation to sit host exams while preparing for home university finals, the Link Tutor facilitated a flexible approach allowing them to prioritise their original degree. This culturally fluent mentorship bridged the gap between the university’s rigid structures and the students’ fluid realities.

#### 4. Sanctuary Institutional Ethos

The overarching culture of the university acted as a profound facilitator. The University established identity as a ‘University of Sanctuary’ provided the moral and administrative vocabulary required to launch the initiative swiftly. Because the cultural groundwork of inclusion and global responsibility was already laid, the proposal of hosting displaced students did not face ideological resistance at the executive level. A senior leader reflected that the school is empowered to act because “*education has to be a beacon of light… and I think institutions such as ours should be doing the right thing”*. This entrenched civic culture meant that arguments regarding risk or resource drain were rapidly superseded by the institution’s self-concept as a global humanitarian actor.

#### 5. Peer-to-Peer Support Systems

Internal student-led resources proved to be an essential facilitator for social and academic survival. The culture of the student body, which actively desired to support the displaced scholars, was leveraged through formal and informal peer networks. Recognising the limits of faculty intervention, the programme arranged “*a peer supporter for them… a student in their year we’ve volunteered”*. This near-peer mentoring helped decode the unspoken rules of the host institution. Across various international contexts, students echoed this necessity. A student relocated to South Africa noted, *“I looked for people like me, being in a group with others who spoke the same language… made adaptation much easier”*. Finding a local cohort that shared study materials and social space was a primary survival mechanism, highlighting the student body as a critical internal resource.

#### 6. Ground-Level Communication Deficits

While high-level strategic alignment was strong, a notable barrier was the cultural disconnect and communication deficit at the frontline clinical level. Teaching faculty and placement coordinators often remained ignorant of the specific allowances granted to the displaced students. A tutor noted *“a lack of information understanding about how the students are expected to attend somewhat not all days… learning providers will look at it that way, that they have been asked to provide so many tutorials… and these students are… hardly here”*. This resulted in friction where displaced students were unfairly perceived as uncommitted or absent, forcing them to constantly self-advocate and correct their instructors. The failure to distribute internal guidance effectively to external clinical partners created unnecessary daily stress.

#### 7. Flexible Placement Allocations

The capacity of the medical school to marshal internal logistical resources to alter standard placement rules was a key facilitator. Because the students were not seeking a UK degree, the school had the flexibility to mitigate stringent placement attendance rules. When one student required a neurosurgery rotation - which is not standard in her host School’s undergraduate curriculum but was required by her home school in Gaza - the host institution utilised its internal networks to *“arrange for her to go on an elective basically to Ireland to do her neurosurgery requirement”*. This internal logistical agility, supported by administrative willingness to bend standard timetabling, was crucial in satisfying the bespoke academic needs of the displaced cohort.

#### 8. Hierarchical Distance in Clinical Settings

A major cultural barrier encountered by students across various global hosting sites was the severe shift in clinical hierarchy and professional interaction. In Gaza, students reported a highly communal, accessible teaching culture. In stark contrast, students relocated to Pakistan and Malaysia faced intense hierarchical rigidity. A student in Pakistan observed, “*Here, you are a visiting student, so there is some degree of indifference from doctors”* and noted that *“students are not allowed to participate in clinical work… honestly, this is very frustrating”*. Similarly, a student in Malaysia observed that *“communication is more formal… Here, before asking a question, you need to check if it is appropriate”*. This vast cultural distance in student-doctor dynamics forced displaced students to suppress their natural learning instincts, creating a profound barrier to clinical integration.

#### 9. Financial Burden Mitigation

The elimination of financial barriers acts as a primary facilitator for programme initiation, as host institutions recognise that displaced students lack the economic infrastructure to support themselves. The analysis indicates that the hosting school’s willingness to participate hinged heavily on internal agreements to absorb baseline costs. For example, the head of accommodation confirmed that *“as part of the [displaced students] initiative, the accommodation fees would be waived, so therefore there’d be no cost to the student,”* a decision directly authorised by university senior management. Furthermore, tuition fees were universally waived by the host universities in the UK, representing a significant opportunity cost that the institution absorbed to facilitate the programme. By removing these baseline financial hurdles, the programme shifted from a commercial transaction to a humanitarian intervention, allowing administrative units to support displaced students without requiring complex financial clearance.

#### 10. Philanthropic Capital Integration

While the university waived internal fees, the successful start of the programme required external financial facilitators to cover day-to-day survival. The integration of philanthropic capital emerged as a crucial feature, as universities often cannot directly fund student living expenses. The analysis highlights that external charities, such as the Hanoon Foundation, which donated £78,000 to cover living costs for two hosted students for the duration of their study, provided the necessary financial scaffolding. Interviewees noted that this *“funding removes at least one of the barriers to the students’ ability to complete their studies”*. Furthermore, securing these funds was deliberately kept separate from the university’s core budget, ensuring that the school executive’s “*primary concern… to not an adversely affect the provision of resource to the existing students”* was respected. This triangulation of university fee waivers and external charitable stipends proved to be a highly effective programme feature.

### Research Question 4: Attitudes and Beliefs of Staff and Faculty

This research question examines the psychological and ideological climate of the hosting institution, analysing how faculty attitudes - including moral imperatives, fears of academic dilution, trauma assumptions, egalitarian blindness, and cultural misinterpretations - impact programme execution.

Each theme generated from the analysis and presented in Table 4 is further interpreted below.

**Table 4.** Staff and Faculty Attitudes Affecting Participation in Hosting Displaced Medical Students.

| Theme | Theme Definition | Corresponding Concepts | Interview No. |
| --- | --- | --- | --- |
| **The Moral Imperative** | The belief that hosting is a fundamental ethical duty transcending administrative hurdles. | Doing the right thing,<br>Social responsibility | Int 10, Int 14 |
| **Fear of Academic Dilution** | Faculty anxiety regarding the academic capability of displaced students and historical failures. | Negative previous experience, Rigour maintenance | Int 8 |
| **Assumptions of Trauma** | Preconceived beliefs about the psychological fragility of displaced individuals. | PTSD assumptions,<br>Over-medicalizing grief | Int 2, Int 4, Int 12 |
| **Egalitarian Blindness** | The belief that treating displaced students exactly like domestic students is the fairest approach. | Colourblind integration,<br>Assuming baseline understanding | Int 11, Int 13 |
| **Empathy vs. Clinical Practicality** | The tension between feeling sorrow for the students and enforcing rigid clinical standards. | Clinical attendance strictness, Systemic rigidity | Int 3, Int 11 |
| **Resistance to Hierarchical Displacement Bias** | The belief that humanitarian aid must be universally applied regardless of geopolitical popularity. | Universal displacement rights, Resisting double standards | Int 1, Int 12 |
| **Trust in Colleague Competence** | The reliance on the expertise of specific faculty members to vouch for the initiative. | Trust in displaced initiative coordinator, Delegated authority | Int 6, Int 14 |
| **Misinterpretation of Cultural Cues** | Faculty misunderstanding of displaced students' communication styles and assertiveness. | Loudness as rudeness, Hesitancy as incompetence, Cultural disconnects | Int 2-6, Int 2-8, Int 4-8 |

#### 1. The Moral Imperative

The most pervasive belief facilitating the acceptance of the programme was the deep-seated conviction among senior staff that hosting the students was an undeniable moral imperative. When asked about their assumptions, faculty consistently bypassed logistical reasoning to focus on ethical duty. A senior executive explicitly stated, *“it’s fundamentally the right thing to do… you just have to stand up and do the right thing sometimes, and society needs a good dose of that”.* This attitude acted as a powerful solvent against bureaucratic resistance; because the initiative of hosting displaced students was framed as a humanitarian necessity in the face of widespread destruction, staff were highly motivated to find solutions rather than highlight obstacles. This shared moral compass aligned disparate departments toward a common goal.

#### 2. Fear of Academic Dilution

Conversely, a significant barrier to acceptance was the historical anxiety among assessment faculty regarding academic dilution. Previous negative experiences heavily coloured faculty attitudes. An assessment lead recounted a *“very difficult experience… when we took in some international students… they performed really badly in the exams… and I didn’t want them [the Gaza students] to be set up for failure”*. This assumption - that transferring students from vastly different pedagogical backgrounds into mid-stream clinical years would result in catastrophic failure - made faculty hesitant to grant full summative integration. It was in part by shifting to formative assessments that this fear was mitigated, protecting both the institution’s academic rigour and the students’ psychological wellbeing from the trauma of potential failure.

#### 3. Assumptions of Trauma

Faculty attitudes regarding the mental state of the displaced students operated as both a facilitator of care and a barrier to agency. Many staff entered the initiative with the assumption that the students would be severely traumatised and require intense psychological management. The programme lead noted, *“there was an expectation that there would be trauma here… and there was a concern or almost a worry as to how would we be able to address… that trauma”*. While this belief facilitated the deployment of pastoral care, it also risked over-medicalising the students’ grief. Recognising this, astute faculty shifted their attitude to be *“guided by the students, by the individuals themselves and just be available… rather than us making assumptions as to 1. whether they need it, 2. how much”*.

#### 4. Egalitarian Blindness

A dominant belief among clinical and frontline teaching staff was that the most respectful way to integrate displaced students was to treat them entirely identically to the UK cohort. A clinical supervisor proudly noted, *“We treat them essentially as a Leicester student… unless we are told they have specific needs”*. While this attitude facilitated a sense of dignity, avoiding the stigma of the refugee label, it inadvertently created barriers. Students often felt lost and unable to ask for help because staff assumed they possessed standard localised knowledge, unaware of their status as displaced students. This left students realising that *“they [the staff] didn’t consider this difference to support”*. One pilot scheme has thus since modified its approach to informing clinical placement leads of these hosted students, and asking the hosted students to likewise introduce themselves to the leads as they rotate to ensure both students and staff feel better supported.

#### 5. Empathy vs. Clinical Practicality

The analysis reveals a tension in staff attitudes between humanitarian empathy and the rigid demands of clinical practicality. While faculty universally expressed sorrow for the displaced students’ circumstances, the operational demands of hospital placements required strict adherence to attendance and performance metrics. Some ground-level educators exhibited frustration when displaced students missed clinical days to study for their home university exams. A personal tutor observed *“a bit of a dichotomy in the minds of the learning providers that, oh, I haven’t been able to provide the students with the same opportunities… because they’re hardly here”*. This demonstrates that while high-level attitudes were highly empathetic, ground-level attitudes were at times strictly utilitarian, creating friction for the students.

#### 6. Resistance to Hierarchical Displacement Bias

A critical belief that facilitated institutional buy-in was the conscious rejection of double standards in humanitarian aid. Faculty were acutely aware of the disparity in the governmental and societal response between the Ukraine conflict and the Gaza conflict. A key assumption explicitly voiced by staff was that *“We cannot say to the displaced students coming from country X that you’re good displaced students and then telling to the other… that you’re bad”*. This commitment to universal equity drove faculty to push the initiative forward precisely *because* they felt external actors were ignoring the crisis. The attitude of rectifying systemic geopolitical bias empowered the staff to view the initiative as an act of institutional justice.

#### 7. Trust in Colleague Competence

The acceptance of the programme by wider faculty was heavily facilitated by a deep-seated trust in the competence of the coordinating individuals. In the absence of a comprehensive institutional blueprint, staff relied on their belief in the lead coordinator’s integrity and expertise. A contributing doctor noted, *“because my connection with him, I probably asked fewer questions because I just know he does things in the right way”*. Similarly, the Head of School operated on the assumption that *“I had people around me who would be very able to navigate the water carefully and all I had to do was empower them”*. This horizontal and vertical trust reduced the need for prolonged deliberation and facilitated efficient decision-making, substantially accelerating the programme’s launch.

#### 8. Misinterpretation of Cultural Cues

A subtle but impactful barrier to the programme’s acceptance on the ward floor was the misinterpretation of cultural cues by host faculty. Displaced students often possessed communication styles that differed from local expectations. A student in Malaysia eloquently highlighted this barrier, noting, *“People here tend to speak softly… Our normal way of speaking can sound loud to them, and the way we interact with each other or with doctors may seem impolite from their perspective… we were often misunderstood at the beginning”*. Furthermore, in Turkey, a student recounted a deeply distressing moment of explicit linguistic rejection when a patient “*told my Turkish colleague that he did not come to be treated by a foreigner”* upon hearing Arabic spoken. Overcoming these entrenched attitudes required immense resilience from the students to recalibrate their intercultural communication expectations.

### Research Question 5: Planning and Leadership Factors

This research question evaluates the operational and strategic leadership dynamics - encompassing scheduling, coordination, bureaucratic navigation, risk mitigation, and top-cover shielding - that facilitated the successful launch of the hosting programme.

Each theme generated from the analysis and presented in Table 5 is further interpreted below.

**Table 5.** Planning and Leadership Factors Affecting Participation in Hosting Displaced Medical Students.

| Theme | Theme Definition | Corresponding Concepts | Interview No. |
| --- | --- | --- | --- |
| **Discretionary Leadership Time** | The allocation of autonomous, unmonitored time by senior faculty to coordinate the complex logistics. | Uncontracted hours,<br>Diary autonomy | Int 2, Int 14 |
| **Agile Implementation Over Perfection** | The leadership decision to launch the programme rapidly despite logistical imperfections to prevent further study loss. | Good enough planning,<br>Iterative development | Int 2, Int 14 |
| **Decentralised Coordination** | Empowering specific champions to manage the programme rather than routing all decisions through centralised committees. | Autonomy, Champion-led execution | Int 2, Int 14 |
| **Strategic**<br>**Administrative**<br>**Flexibility and**<br>**Framework**<br>**Repurposing** | Planning the initiative around existing administrative categories and established institutional pathways, enabling implementation within prevailing regulatory and governance frameworks rather than creating new administrative mechanisms. | Extended elective status, Visiting student classification, Alternative enrolment pathways, Working within GMC-approved frameworks | Int 2, Int 5, Int 11, Int 12, Int 14 |
| **Long-Term**<br>**Scalability**<br>**Vision** | Planning the pilot not just as a one-off rescue, but as a blueprint for a sustainable, global institutional response to displacement. | Establishing networks, Future crisis readiness | Int 2, Int 14 |
| **Triangulating**<br>**Stakeholder**<br>**Needs** | The leadership task of balancing the demands of the host university, home university, and the students. | Flexible partnership mechanisms, Aligning dual curriculums | Int 2, Int 3, Int 6 |
| **Mitigation of**<br>**Institutional**<br>**Risk** | Planning factors designed to protect the host university from legal, financial, and academic liabilities. | Non-summative exams, External charity funding | Int 8, Int 14 |
| **Protective**<br>**Leadership**<br>**Shielding** | The role of senior leadership in absorbing institutional friction to protect the operational team. | Executive top-cover, Using strong performance metrics | Int 14 |
| **Beneficiary-Led Agency** | Planning the programme around the dignity and specific career trajectories of the beneficiaries rather than donor demands. | Beneficiary-led KPIs, Preserving home ties | Int 4, Int 10 |

#### 1. Discretionary Leadership Time

The most vital internal resource facilitating the programme was the discretionary time and emotional labour invested by specific clinical leaders. Without dedicated champions, the bureaucratic inertia of the university might have acted as an impenetrable barrier. The Head of School explicitly highlighted this, noting that the primary driver was a specific professor’s time, stating he *“spends many, many more hours working than he is contracted to… he has autonomy over his diary… and I was very happy to trust him”*. This autonomy allowed the lead coordinator to navigate the intricate logistics of curriculum mapping, charity funding, and pastoral care. However, the analysis also notes the danger of this reliance; relying heavily on the *“emotional energy that goes into it”* creates sustainability risks if that individual were to leave or experience burnout.

#### 2. Agile Implementation Over Perfection

A critical leadership factor facilitating the rapid deployment of the programme was the conscious rejection of bureaucratic perfectionism. Given that the displaced students were actively losing academic years, the programme lead adopted an agile implementation strategy. He explicitly noted the danger of *“making the good the enemy of the perfect… I didn’t want to wait till we had everything lined up to say, OK, now we’re ready to receive you because that would be another year or two years… lost for these students”*. This planning philosophy accepted that gaps, such as imperfect pastoral alignment or placement confusion, would occur, but prioritised the immediate resumption of study. This bias toward action over endless committee deliberation was essential for the successful start of the initiative.

#### 3. Decentralised Coordination

The successful initiation of the programme relied heavily on decentralised leadership. Rather than managing the crisis response through a monolithic central university task force, the medical school empowered a specific clinical academic to act as the autonomous focal point. The Head of School described his leadership role not as a micro-manager, but as a facilitator of autonomy: *“I was very happy to trust him to use his time to best effect and certainly would not have been seeking to get him to account for his time”*. This Decentralised approach allowed the lead coordinator to act swiftly, negotiating directly with external charities, home institution deans, and clinical placement leads, keeping the School leadership up to date, without requiring multi-tiered approval for every micro-decision.

#### 4. Strategic Administrative Flexibility and Framework Repurposing

Perhaps the most ingenious planning factor was the strategic repurposing of existing administrative categories to work within strict governmental and institutional quotas on medical school admissions. Creating a novel legal pathway for displaced medical students would have triggered insurmountable barriers with the UK Visas and Immigration (UKVI) office, the General Medical Council (GMC), and university admissions caps for international medical students. Enrolling new full-time students was impossible, because medical placements are heavily capped. Leadership addressed this by planning the initiative as an extended elective, study abroad, or visiting student programme – as these hosted students ultimately remained students of their home school from which they have been displaced (a key aspect of this initiative to also support at-risk institutional continuity) A strategic leader explained, *“Because we already have study abroad style schemes… why not apply it to this?”*. He further noted that by framing it this way, *“a whole raft of regulations within higher education institutions… completely dissolve”.* This agile strategic and administrative manoeuvring, working within already existing systems and pathways, allowed the university to integrate the students legally and provide them with educational resources without violating strict national medical enrolment quotas, serving as a masterclass in bureaucratic navigation.

#### 5. Long-Term Scalability Vision

While the immediate goal was to rescue the educational trajectories of specific students, the planning was profoundly facilitated by a vision of long-term scalability. Leadership did not want to reinvent the wheel for the next geopolitical crisis. The goal was to ensure *“this becomes institutionalised… such that we are able to respond at the point of need… completely proactively, rather than in the way that it has been done in the past reactively”*. By documenting the process, presenting to the Medical Schools Council, and sharing the blueprint with all UK medical schools, the leadership transformed a localised pilot into a scalable national model. This visionary planning justified the heavy initial investment of time and resources to the university executive.

#### 6. Triangulating Stakeholder Needs

A major planning barrier was the necessity of triangulating the conflicting needs of three distinct entities: the host university, the displaced students, and the devastated home university in Gaza. Leadership had to ensure that the intervention did not inadvertently strip the home university of its students and revenue. A coordinator from Gaza noted the challenge of managing *“challenges on the curriculum and in the… financial problem”* without a formal MoU. Leadership navigated this by ensuring the students remained enrolled in their home institutions and tailoring the host experience to generate evidence for the home dean’s progression boards. This delicate diplomatic planning preserved the sovereignty of the conflict-affected institution while providing the physical safety of the host university.

#### 7. Mitigation of Institutional Risk

University executives are understandably inherently risk-averse, and a key leadership factor in gaining approval was the proactive mitigation of institutional risk. Planning was meticulous in ensuring the initiative cost the university very little in hard currency, leveraging external donors for living costs. Furthermore, by establishing that the students would take exams formatively and would not be awarded a degree by the host institution, leadership removed the risk of academic liability and regulatory censure. As the Head of School noted, *“I only believe in asking for things… that I think we have a reasonable chance of getting… Would we be creating precedence the university couldn’t live up to?”*. By designing a low-cost, low-liability, high-impact programme, leadership effectively neutralised internal administrative resistance.

#### 8. Protective Leadership Shielding

The operational success of the initiative was heavily dependent on the support and guidance provided by senior leadership. The Head of School utilised the medical school’s strong performance metrics (such as top National Student Survey scores) as political capital to encourage support for the initiative from central university. He noted, *“things like the NSS score empower us to do things like this… we’re given the benefit of the doubt where uncertainty exists from other quarters”.* By projecting confidence and absorbing any political or administrative friction generated by the central university, senior leadership created a safe operational bubble wherein the project champions could execute the logistics of the programme with wider institutional buy-in and support.

#### 9. Beneficiary-Led Agency

Finally, the planning phase was heavily guided by a commitment to student-centred agency, deliberately avoiding the saviour complex that often plagues humanitarian interventions in higher education. Leadership planned the programme so that students *“weren’t made to feel that they somehow owe someone something… that they have a right to be here, that they’re not second class citizens”*. Planners resisted the urge to use the students for donor publicity, ensuring KPIs were beneficiary led rather than donor-driven. From the students’ perspective, navigating the lack of formal support structures birthed a relentless internal agency. A student in Pakistan succinctly defined their survival planning: *“’Survival of the adaptable’ is the principle I learned the most after leaving Gaza… you must expect that you cannot give 100% to all aspects at the same time”.* Ultimately, this widespread individual perseverance, when successfully paired with flexible institutional accommodations, became the primary driver of the students’ academic continuity.

## Discussion

To contextualise these qualitative findings and extract their broader scientific value, this section translates the empirical results through the theoretical lens of the Consolidated Framework for Implementation Research (CFIR), details how these findings theoretically advance the framework, outlines practical and policy implications, and delineates study limitations alongside future research directions.

### Theoretical Interpretation through the CFIR

Analysing the qualitative findings through the lens of the CFIR reveals how multi-level determinants interact to shape the trajectory of this public sector innovation. Within the Innovation Characteristics domain, the structural decision to implement *Formative Assessment Adaptation* operated as a decisive facilitator. By altering the trialability and complexity of academic progression and decoupling host feedback from degree-awarding mechanisms, the programme significantly reduced the perceived risk for conservative institutional decision-makers. However, this adaptation created a massive tension in compatibility for the students, who were forced to engage in the exhausting, simultaneous negotiation of a *Dual-Curriculum*. This conflict demonstrates that an innovation feature designed to facilitate organisational adoption can simultaneously introduce severe operational friction for the end-recipients. One at-risk institution has addressed this through accrediting hosted students’ learning through their host institution – hosted (i.e. Gazan) students learn and are assessed entirely with host (e.g. UK) students, and submit their results for accreditation and progression/award decisions by their home university. This ensures complete alignment of hosted student learning with assessment, which becomes much more challenging if there as ongoing requirement for hosted students to sit their home school’s written exams remotely.

In the Outer Setting domain, we observe a highly hostile, restrictive context characterised by *Hostile Immigration Frameworks* and rigid *Medical Regulatory Compliance*. Traditional implementation science often conceptualises the Outer Setting as a static environment of policies and mandates [13]. Here, however, the Outer Setting operated as an active, aggressive force of systemic resistance. The UKVI’s border controls and The Office for Students’ placement caps represented powerful structural barriers that threatened to completely halt the initiative. Interestingly, the data reveals that this external hostility triggered a powerful ‘tension for change’ within the Inner Setting. The absolute contrast between governmental support for Ukrainian scholars and the *Geopolitical and Governmental Inaction* regarding Gaza catalysed a profound moral response within the university.

This moral response was structurally supported by the university’s pre-existing *Sanctuary Institutional Ethos*. Within CFIR, culture and compatibility are recognised as critical Inner Setting determinants [13]. Because the host institution possessed a deeply embedded civic culture that prioritised global social responsibility, the hosting initiative was highly compatible with the school’s self-concept, effectively neutralising executive-level anxieties regarding financial cost or reputational backlash. However, a critical ‘networks and communication’ barrier emerged as a *Ground-Level Communication Deficit*. While strategic leaders established flexible placement policies, the failure to transmit this guidance to frontline clinical providers resulted in occasional friction on the ward floor, where students were unfairly stigmatised for dual-curriculum absences. This highlights that high-level cultural alignment in the Inner Setting does not automatically translate into effective implementation climate on the frontline.

This clinical friction was successfully mediated by determinants in the Characteristics of Individuals domain, most notably the *Moral Imperative* voiced by senior staff and the deployment of *Cultural Translators and Link Tutors*. The Link Tutor acted as a vital boundary-spanning role, leveraging high self-efficacy and cultural fluency to bridge the academic gap between the host and home environments. This agential intervention was reinforced by *Peer-to-Peer Support Systems*, where domestic student cohorts actively mobilised to provide informal academic and social orientation.

Finally, within the Implementation Process domain, the success of the initiative was driven by *Decentralised Coordination* and *Agile Implementation Over Perfection*. After initial university level approval, rather than waiting for consensus across slow-moving central university committees for each decision, the Head of School delegated substantial authority to a dedicated project champion. This champion utilised Strategic Administrative Flexibility, drawing on existing visiting-student and elective pathways to integrate students within established institutional and regulatory frameworks while avoiding the need to create entirely new administrative mechanisms. This process was supported by central university through Protective Leadership Shielding, where the Head of School utilised the department’s elite performance metrics (such as top NSS scores) as political capital to absorb administrative friction, creating a safe operational space for rapid execution.

### Advancing the CFIR

The ultimate value of this study lies in its capacity to theoretically advance the Consolidated Framework for Implementation Research (CFIR), extending its utility from stable, high-resource clinical environments to highly dynamic, politically contested, and crisis-driven public sector environments. When applied to rapid humanitarian interventions within public institutions, traditional CFIR exhibits two major theoretical limitations that this study directly addresses.

We have conceptualised these additions visually in Figure 1, demonstrating how active resistance, leadership shielding, and systemic boundary-crossing restructure the passive concentric settings of the traditional framework.

**Figure 1.**
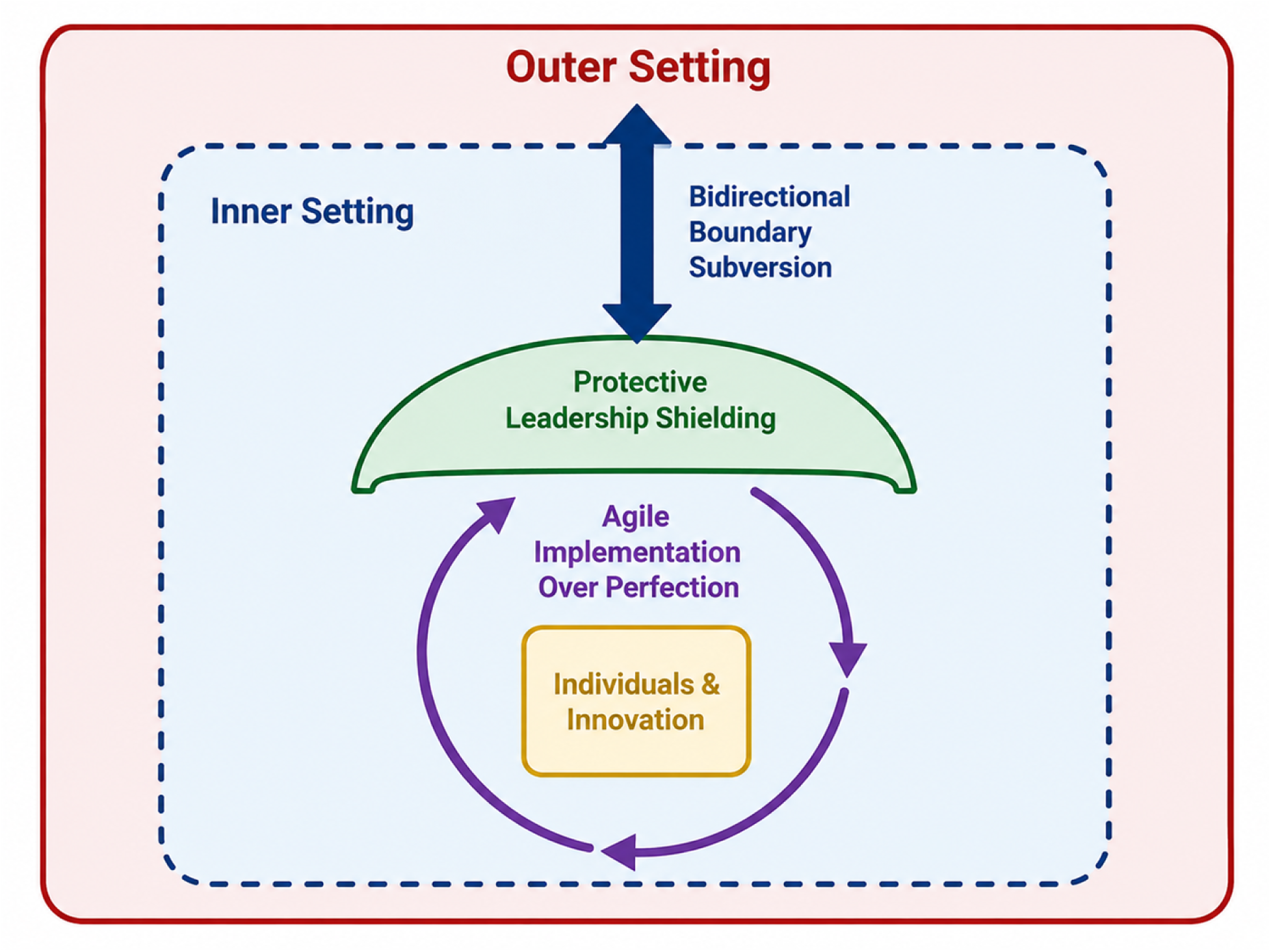
The Advanced, Crisis-Adapted CFIR

First, CFIR has historically conceptualised the Outer Setting as a passive, macro-level context that indirectly influences the Inner Setting through policies or peer pressure [13]. However, this study demonstrates that in crisis-driven equity interventions, the boundary between the Outer and Inner Settings is highly active, morally charged, and bidirectional. We propose the introduction of ‘Bidirectional Boundary Subversion’ as a formal construct within the Outer/Inner Setting interface. This construct captures the process where grassroots institutional actors do not merely react to external policy constraints (such as hostile immigration laws or regulatory caps) but actively identify internal administrative gaps (such as through framework repurposing) to subvert and push back against, yet still work within, those external policies. In this model, the Inner Setting’s moral ethos actively resists the Outer Setting’s political apathy, transforming the public institution from a passive implementer of state policy into an active global ethical agent.

Second, traditional CFIR processes are structured around highly linear, planned, and systematically evaluated phases of execution [16]. In a rapid humanitarian crisis, waiting for perfect administrative alignment is a recipe for complete failure, resulting in the prolonged academic stagnation of vulnerable populations [2]. To address this, we propose the integration of ‘Agile Implementation Over Perfection’ as an active construct under the Implementation Process domain. This construct theorises that in crisis environments, successful implementation requires a deliberate, strategic rejection of bureaucratic perfectionism, prioritising the immediate resumption of study and accepting that operational gaps must be resolved iteratively on the ground. Concurrently, we propose the addition of ‘Protective Leadership Shielding’ as a formal construct under the Inner Setting / Leadership Engagement domain. This construct captures the process where senior executives do not merely support an innovation but actively utilise their accrued political capital and performance metrics to absorb organisational friction and shield project champions by engaging central administrative structures. This creates a safe, decentralised operational bubble that is essential for rapid, high-risk equity interventions.

### Theoretical and Practical Implications

At the theoretical level, this study reframes how educational hosting programmes for displaced scholars are conceptualised in the literature. Rather than viewing these programmes as simple, reactive, and short-term individual rescue missions, this paper demonstrates that they operate as vital instruments of inter-institutional preservation and global justice. When conflict-zone medical schools face scholasticide, the global higher education system can act as a decentralised, moral safety net [2]. Furthermore, by integrating a critical realist ontology with implementation science, this study proves that structural barriers are not absolute; rather, they exist alongside powerful, localised agential strivings that possess the capacity to exploit administrative grey areas and subvert hostile state policies.

At the practical level, this study provides a highly replicable, structural blueprint for medical schools and higher education institutions globally to manage educational sanctuary initiatives with high structural feasibility and localised academic safety. Crucially, institutions must enact a strategy of formative decoupling by designing academic pathways strictly around non-summative, formative-only clinical assessments and qualitative feedback portfolios. This tactical adaptation successfully shields the hosting institution from student progression, graduation, and degree-awarding liabilities, while simultaneously furnishing the displaced students with the rigorous, accredited evidence required by their original home universities to document continued progress. To overcome the significant barriers presented by rigid national enrolment quotas, visa limits, and immigration hurdles, leadership teams should utilise a mechanism of administrative framework repurposing. This is achieved by registering hosted cohorts under pre-existing, low-friction administrative categories, such as ‘extended clinical electives’ or ‘visiting scholars / Study Abroad schemes’ rather than trying to establish novel legal enrolment pathways or matriculating them as standard full-time students (the latter of which also ends the connection between the student and their home at-risk institution).

Additionally, successful integration requires dedicated resource provision, most notably the formal appointment of culturally fluent ‘Link Tutors’ who act as academic and pastoral translators. These specialised mentors navigate the structural tension of dual-curriculum balance and mitigate the communication disconnect frequently encountered between strategic deans and frontline clinical supervisors. To ensure the financial feasibility of the initiative, universities must strategically triangulate capital by combining institutional tuition and housing fee waivers with external, philanthropic funding. Securing independent, third-party stipends to cover day-to-day living expenses, and managing these funds completely outside the core institutional budget, enables the project to remain cost-neutral, effectively neutralising executive-level anxieties regarding the diversion of public resources from domestic cohorts. Finally, institutions must drive sector-wide diffusion by proactively sharing their operational templates and clinical mapping protocols with national medical school councils. This collective collaboration helps to establish a highly coordinated, multi-site network of sanctuary, distributing administrative risk, validating standard operating procedures, and scaling the hosting template to prepare for future global educational crises.

### Limitations and Suggestions for Future Research

While this study offers a highly detailed, theoretically advanced qualitative analysis, several limitations must be acknowledged. First, the qualitative dataset, representing 66 in-depth interviews, is concentrated within specific international host sites, with the UK serving as a primary context. While the multi-site design captures diverse realities in Malaysia, Pakistan, Turkey, and South Africa, the findings may be shaped by the unique resources and legal frameworks of these specific nations. Second, this research relied entirely on qualitative, interview-based data. It did not incorporate longitudinal quantitative tracking of the students’ clinical competency development, academic test scores, or long-term career progression after graduation. Third, because the programme was rapidly implemented during an active geopolitical crisis, there is a risk of recall bias among organisers, and accessing the most vulnerable, unregistered displaced students who fled to transit nations remained extremely difficult. Finally, the study focuses on displacement arising from a single conflict context. While this focus enabled an in-depth examination of the mechanisms supporting continuity of medical education during crisis, the transferability of the findings to students displaced by other conflicts, political crises, or humanitarian emergencies remains to be established.

To address these limitations, future research should employ longitudinal, mixed-methods designs. Researchers should quantitatively evaluate the clinical competency trajectories and professional licensing pass rates of hosted displaced students over a multi-year period, comparing their outcomes with domestic cohorts. Furthermore, future studies should test the applicability of this crisis-adapted CFIR framework across other highly regulated public sector professional training domains - such as nursing, pharmacy, dentistry, and engineering - that are experiencing systemic displacement due to ongoing global crises. Finally, comparative policy analyses should investigate how different national visa and border control frameworks across Europe, North America, and the Global South systematically facilitate or block the execution of academic sanctuary initiatives.

## Conclusion

This multi-national study demonstrates that hosting displaced medical students is a highly complex, multi-systemic educational intervention that cannot be understood or executed through traditional, static administrative structures. The qualitative findings reveal that the ultimate success of these humanitarian initiatives is not determined by a single factor, but is shaped by the dynamic, non-linear interaction between structural programme features (such as formative assessment adaptation and dual-curriculum flexibility), hostile external forces (such as immigration barriers and clinical caps), and powerful internal facilitators (such as a strong sanctuary institutional ethos, peer networks, and dedicated Link Tutors).

The final, synthesised message of this study is that higher education institutions must reject the passive, risk-averse saviour complex that has historically characterised refugee educational support. Hosting displaced cohorts must not be conceptualised as a temporary, sentimental act of individual charity, but rather as an act of global systemic resilience and institutional preservation. When local healthcare training pipelines in conflict zones face systematic destruction, the global higher education system possesses the capacity to operate as a laminated, decentralised moral safety net [2]. By utilising strategic administrative flexibility and decentralised coordination, academic deans and clinical leaders can successfully subvert and push back against hostile national policies, proving that public institutions can act as powerful, independent stewards of global social justice and public health.

This systemic transformation is theoretically illuminated when evaluated through Joseph Schumpeter’s innovation framework, which conceptualises the initiative as a radical public sector educational disruption across five key dimensions [22]. First, as a new good (or product), the programme introduces a risk-neutral, formative-only assessment pathway that successfully decouples degree-awarding legal liabilities from clinical training validation. Second, the new method of production establishes a hybrid, dual-stream pedagogical mapping process, facilitating simultaneous clinical-placement engagement and virtual home-curriculum tracking through bespoke IT provisions and Link Tutors. Third, it opens a new avenue for participation by utilising existing educational pathways and administrative mechanisms within national regulatory frameworks, thereby creating dedicated opportunities for under-represented and highly vulnerable displaced student cohorts. Fourth, a new supply source of capital is secured through the strategic triangulation of cost-neutral university tuition and housing waivers with independent, philanthropic stipends, eliminating direct municipal financial strain on host budgets. Finally, the programme creates a new organisational model by establishing a decentralised, trust-based collaborative network that complements formal institutional arrangements through agile, dean-level coordination supported by national medical councils. Together, these five dimensions embody a process of educational creative destruction, proving that public universities can act not merely as static educational providers, but as dynamic, agile social enterprises capable of spearheading systemic global sanctuary networks.

## Statements and Declarations

### Ethics Approval

This study was formally reviewed and approved by the Institutional Research Ethics Committee and all procedures were conducted in strict accordance with the ethical standards of the institutional research committee and the 1964 Declaration of Helsinki and its later amendments.

### Consent for Human Participants

Prior to their inclusion, written informed consent was obtained from all participants, who were fully briefed on the study’s objectives and their right to voluntary withdrawal at any stage without penalty. To safeguard participant privacy, all data were rigorously anonymised during the analytical phase.

### Availability of data and material

The datasets generated and analysed during the current study are available and will be provided upon a reasonable request.

### Declaration of Generative AI Use

During the preparation of this work, the authors used Gemini to improve readability and assist with language editing. After using this service, the authors reviewed and edited the content as needed and take full responsibility for the final content of the publication.

